# Anthropometric measurements as predictors of hypertension in Busia, Vihiga, Trans Nzoia and Siaya counties of Western Kenya

**DOI:** 10.1101/2021.07.07.21258941

**Authors:** Thomas Andale, Vitalis Orango, Gerald O. Lwande, Grace W. Mwaura, Richard Mugo, Obed K. Limo, Ann Mwangi, Jemima H. Kamano

## Abstract

Emerging data suggest a rise in the incidence rate of hypertension in many countries within Sub-Saharan Africa. This has been attributed to socioeconomic factors that have influenced diet and reduced physical activity further deranging anthropometric measurements. We assessed the predictive power of three anthropometric indicators namely: waist circumference (WC), waist to height ratio (WHtR) and body mass index (BMI) in detecting hypertension. This cross-sectional community survey was conducted in four counties within Western Kenya between October 2018 to April 2019 among 3594 adults. The participants’ sociodemographic data were collected using an interviewer-administered questionnaire and anthropometric measurements taken. We used the R-software for descriptive and inferential statistical analysis. Pearson chi-square test was used to assess the association between anthropometric measurements and hypertension while logistic regressions estimated the likelihood of hypertension. Youden method was used to identify optimal anthropometric cut-offs for sensitivity, specificity and area under the receiver operating characteristics (ROC) curve computation. The crude prevalence of hypertension was 23.3%, however it rose with advancement in age. Furthermore, obese individuals had a three-fold (AOR=2.64; 95% CI: 2.09, 3.35) increased likelihood of hypertension compared to those with a normal BMI. The optimal WC cut-off was 82.5cm for men and 87cm for women, an optimal WHtR of 0.47 for men and 0.55 for women; while the optimal BMI cut-off was 23.7 kg/m^2^ and 22.6 kg/m^2^ for men and women respectively. The sensitivity of WC, WHtR and BMI for men was 0.60, 0.65 and 0.39 respectively and 0.71, 0.65 and 0.78 respectively for women. BMI is the best predictor for hypertension among women but a poor predictor for men; WC had a high hypertension predictive power for both gender while WHtR is the best hypertension predictor for men.

## Introduction

Hypertension as a non-communicable disease of the cardiovascular system has been reported to be a major cause of morbidity and mortality in many countries around the world [1–3]. Hypertension increases the risk of cardiovascular and coronary heart disease [4]. Previous studies have indicated that risk factor modification could help bend the rising trend of the disease [4–6]. This is despite the anecdotal data reporting higher hypertension prevalence rates in countries with developed economies and higher per-capita income [7,8]; most of which are located within the Mediterranean and Atlantic regions [9–11]. However, emerging data from low and middle income countries such as those in Sub-Saharan Africa have reported rising incidence rates for hypertension [6]. This has been attributed to sedentary lifestyle and dietary changes which directly affect the anthropometric characteristics of the affected individuals [5,12,13]. Anthropometric measurements such as body-mass index (BMI), waist circumference (WC) and waist to height ratio (WHtR) are simple methods to assess body composition [5,12,14]. They describe the body’s size, shape, mass and level of fatness as an initial screening technique for obesity and its associated complications such as hypertension. Although the World Health Organisation (WHO) instituted BMI as an epidemiological technique to assess obesity, it does not account for body-fat distribution. On the other hand, WC and WHtR have been reported as optimal techniques to describe more body fat, abdominal and central obesity as well as estimation of visceral adipose tissue that are often associated with metabolic changes that could result in hypertension. This association between anthropometric indices and hypertension have been previously reported in studies conducted in various countries across the globe with differing ethnicities and ancestry [13,15–17]. However, all these studies have reported varying optimal anthropometric techniques and cut-offs for predicting hypertension in the targeted populations [7,13,15,16]. For instance, among elderly men in Taiwan [7], Barbados and Cuba [18], WHtR was reported as the best predictor of hypertension. However, in the Japanese [8] and Cuban [18] populations, BMI alone had a higher predictive power for the presence of hypertension; while WC was a better predictor of hypertension among Taiwanese women [7] as well as Greek men and women [19]. Because of this conflicting evidence, it can be inferred that the predictive power of various anthropometric measurements largely depends on the target populations. This study therefore adds to the existing body of knowledge by assessing the predictive power of the three anthropometric indices described by targeting rural and peri-urban dwelling adults of both gender within their communities in Western Kenya.

Although studies that assess anthropometric predictors of cardiovascular diseases such as hypertension have been previously published, most of these have been in controlled environments such as clinical settings and this could bias the reported outcomes [20,21]. Furthermore, most of them have been conducted in other continents with different socio-demographic and economic characteristics for the enrolled study participants making them not spatially generalizable.

## Materials and methods

This was a cross-sectional community survey conducted in four (Busia, Vihiga, Siaya and Trans Nzoia) counties of western Kenya between October 2018 and April 2019. The study targeted adults aged 18 years or more living in the selected counties who were stratified by their county of residence and randomly sampled systematically until a desired sample size of 3,594 was achieved. The sample size was obtained using Fischer’s formula and proportionately distributed to the four counties (Busia = 900; Trans-Nzoia = 897; Siaya= 897 and Vihiga=900). Written informed consent was obtained from all the potential participants prior to enrollment. Interviewer administered questionnaires were conducted at the homes of the study participants to obtain their sociodemographic characteristics (age, gender, marital status, tribe, education level and occupation).

Seated blood pressure measurements were taken using a calibrated Omron M2 compact upper arm blood pressure monitor (Omron Healthcare, Inc., 1200 Lakeside Drive, Bannockburn, Illinois 60015) after the participant had rested for fifteen minutes. The choice of the appropriate blood-pressure equipment was informed by the participant’s cuff size. Three blood pressure readings were then taken after 5-minute intervals and the average reading computed. Hypertension was defined as either a systolic BP>140 mmHg and/or diastolic BP > 90 mmHg. This was followed by weight and height measurements using calibrated digital weighing scales and meter-rule. The BMI was computed by dividing the weight (in kilograms) and height (in squared meters. Based on the BMI values, clients were categorized into Underweight (<18.5 Kg/m^2^) Normal (18.5-24.9 Kg/m^2^), Overweight (25-29.9 Kg/m^2^) and Obese (≥30 Kg/m^2^). Waist-circumference was measured using a tape-measure (calibrated in centimeters) to the nearest whole number. The normal cut-off for males and females was ≤102 cm and ≤88 cm respectively. Abnormal waist circumference values were set at >102 cm for males and > 88 cm for females. Lastly, WHtR was computed by dividing the waist circumference by height in centimeters. Participants with a WHtR ≥0.5 were considered to have a high risk of hypertension, while those below 0.5 had a low risk.

Statistical analyses were performed using R software. The mean and the standard deviation were used to describe continuous data while the count and the percentage were used to describe categorical data. Association between categorical variables were assessed using the chi square test. Group mean differences were tested using analysis of variance (ANOVA) technique. Pearson correlation was used to test for the strength of association between continuous BMI, WC and Weight height ratio. Logistic regressions were used to estimate the odds of hypertension among persons with different categories of anthropometrics levels. Youden method to was used to determine the optimal anthropometric cut-offs. The obtained cut-offs were compared with the recommended anthropometric cut-offs by the World Health Organization in terms of sensitivity, specificity and area under the curve. Ethical approvals were obtained from the Institutional Research and Ethics Committee of Moi Teaching and Referral Hospital and Moi University School of Medicine (approval number 0002090), as well as the National Commission for Science, Technology and Innovation (NACOSTI) (approval number NACOSTI/P/18/74238/24329) and the respective four County Health Management Teams (CHMTs).

## Results

This study enrolled 3,599 community members in four counties within Western Kenya with a nearly equal proportionate representation and gender distribution. The overall mean age of the study participants was 43.85 (± 15.93) years with the youngest population being those from Siaya county with a mean age of 40.76 (± 14.84) years. The difference in mean age distribution (p<0.001) and age categories (p<0.001) was significant across the counties sampled. The mean BMI of all the study participants was 23.83 (± 9.62) kg/m^2^. There was no statistically significant difference in mean and categorized BMI scores as well as weight to height ratios across the counties. Almost three-quarters (n=2634) of the participants enrolled had a normal waist circumference with an overall mean waist circumference of 85.37 (±12.6) centimeters. Hypertension was present among 23.2% (n=836) of all the study participants enrolled with a statistically significant (p< 0.001) difference in distribution across all the counties sampled. (Table 1).

**Table 1:** Summary of Participant characteristics.

| Variable name | Levels | Total<br>N=3575<br>n(%) | Busia<br>N=901 | Trans<br>Nzoia<br>N=888 | Vihiga<br>N=896 | Siaya<br>N=890 | p-value |
| --- | --- | --- | --- | --- | --- | --- | --- |
| **Gender** | Male | 1737 (48.6) | 446 (49.5) | 437 (49.2) | 398 (44.4) | 456 (51.2) | 0.027 |
|  | Female | 1838 (51.4) | 455 (50.5) | 451 (50.8) | 498 (55.6) | 434 (48.8) |  |
| **Age (years)** | Mean (SD) | 43.78<br>(15.91) | 43.16<br>(16.53) | 47.23<br>(16.91) | 44.07<br>(14.59) | 40.68<br>(14.81) | <0.001 |
| **Age Group**<br>**(years)** | 18-29 | 810 (22.7) | 212 (23.5) | 164 (18.5) | 182 (20.3) | 252 (28.3) | <0.001 |
|  | 30-44 | 1097 (30.7) | 302 (33.5) | 253 (28.5) | 258 (28.8) | 284 (31.9) |  |
|  | 45-59 | 982 (27.5) | 226 (25.1) | 256 (28.8) | 276 (30.8) | 224 (25.2) |  |
|  | 60-74 | 576 (16.1) | 117 (13) | 150 (16.9) | 180 (20.1) | 129 (14.5) |  |
|  | 75+ | 110 (3.1) | 44 (4.9) | 65 (7.3) | 0 (0) | 1 (0.1) |  |
| **BMI**<br>**(Continuous)** | Mean (SD) | 23.45<br>(4.74) | 23.59<br>(4.84) | 23.68<br>(4.69) | 23.53<br>(4.91) | 23 (4.48) | 0.011 |
| **BMI**<br>**Category** | Underweight | 534 (14.9) | 136 (15.1) | 119 (13.4) | 142 (15.8) | 137 (15.4) | 0.037 |
|  | Normal | 1979 (55.4) | 488 (54.2) | 493 (55.5) | 470 (52.5) | 528 (59.3) |  |
|  | Overweight | 710 (19.9) | 176 (19.5) | 183 (20.6) | 191 (21.3) | 160 (18) |  |
|  | Obese | 352 (9.8) | 101 (11.2) | 93 (10.5) | 93 (10.4) | 65 (7.3) |  |
| **WC**<br>**(Continuous)** | Mean (SD) | 85.15<br>(11.86) | 84.42<br>(11.89) | 84.98<br>(11.67) | 86.39<br>(12.39) | 84.82<br>(11.39) | 0.003 |
| **WC**<br>**(Categorized)** | Abnormal | 946 (26.5) | 235 (26.1) | 225 (25.3) | 271 (30.2) | 215 (24.2) | 0.023 |
|  | Normal | 2629 (73.5) | 666 (73.9) | 663 (74.7) | 625 (69.8) | 675 (75.8) |  |
| **WHtR**<br>**(Continuous)** | Mean (SD) | 0.51 (0.08) | 0.51<br>(0.08) | 0.51<br>(0.08) | 0.52<br>(0.08) | 0.51 (0.07) | 0.001 |
| **WHtR**<br>**(Categorized)** | High Risk | 1739 (48.6) | 425 (47.2) | 432 (48.6) | 482 (53.8) | 400 (44.9) | 0.001 |
|  | Low Risk | 1836 (51.4) | 476 (52.8) | 456 (51.4) | 414 (46.2) | 490 (55.1) |  |
| **Hypertension** | No | 2752 (77.0) | 693 (76.9) | 590 (66.4) | 688 (76.8) | 781 (87.8) | <0.001 |
|  | Yes | 823 (23.0) | 208 (23.1) | 298 (33.6) | 208 (23.2) | 109 (2.2) |  |

**Legend:** *Abnormal Waist Circumference cut-offs – Men (> 102 cm), Female (>88 cm)*
*Normal Waist Circumference cut-offs - Men (≤102 cm), Female (≤88 cm)*
*WHtR ≥0.5 (High Risk); <0.5 (Low Risk)*

There was no statistically significant difference in the crude hypertension prevalence across gender. Notably the prevalence of hypertension increased with age (p<0.001) with the mean age of those with hypertension being higher at 53.2 compared to 41years (p<0.001). In addition, the prevalence of hypertension significantly increased with increase in BMI (p<0.001), waist circumference (p<0.001) and waist to height ratio (p>0.001) (Table 2).

**Table 2:** **Prevalence of hypertension by sociodemographic factors and anthropometric measurements**

| **Variable name** | **Levels** | **Normal**<br>**2752 (77.0%)**<br>**n (%)** | **Hypertensive**<br>**823 (23.0%)**<br>**n (%)** | **Total**<br>**N=3575**<br>**n (%)** | **p-value** |
| --- | --- | --- | --- | --- | --- |
| **Gender** | Male | 1332 (76.68) | 405 (23.32) | 1737 (48.59) | 0.691 |
|  | Female | 1420 (77.26) | 418 (22.74) | 1838 (51.41) |  |
| **Age (years)** | Mean (SD) | 41 ($\pm 15.2$) | 53.2 ($\pm 14.7$) | 43.8 ($\pm 15.9$) | <0.001 |
| **Age Groups**<br>**(years)** | 18-29 | 758 (93.58) | 52 (6.42) | 810 (22.66) | <0.001 |
|  | 30-44 | 935 (85.23) | 162 (14.77) | 1097 (30.69) |  |
|  | 45-59 | 659 (67.11) | 323 (32.89) | 982 (27.47) |  |
|  | 60-74 | 345 (59.9) | 231 (40.1) | 576 (16.11) |  |
| | $\geq 75$ | 55 (50.0) | 55 (50.0) | 110 (3.08) | |
| **BMI** | Mean (SD) | 23 ($\pm 4.5$) | 24.9 ($\pm 5.3$) | 23.5 ($\pm 4.7$) | <0.001 |
| **BMI**<br>**(Categorized)** | Underweight | 444 (83.15) | 90 (16.85) | 534 (14.94) | <0.001 |
|  | Normal | 1590 (80.34) | 389 (19.66) | 1979 (55.36) |  |
|  | Overweight | 502 (70.7) | 208 (29.3) | 710 (19.86) |  |
|  | Obese | 216 (61.36) | 136 (38.64) | 352 (9.85) |  |
| **WC**<br>**(Continuous)** | Mean (SD) | 83.5 ($\pm 10.9$) | 90.7 ($\pm 13.1$) | 85.2 ($\pm 11.9$) | <0.001 |
| **WC**<br>**(Categorized)** | Abnormal | 619 (65.43) | 327 (34.57) | 946 (26.46) | <0.001 |
|  | Normal | 2133 (81.13) | 496 (18.87) | 2629 (73.54) |  |
|  | Normal | 2553 (78.48) | 700 (21.52) | 3253 (90.99) |  |
| **WHtR**<br>**(Continuous)** | Mean (SD) | 0.5 ($\pm 0.1$) | 0.5 ($\pm 0.1$) | 0.5 ($\pm 0.1$) | <0.001 |
| **WHtR**<br>**(Categorical)** | High Risk | 1197 (68.83) | 542 (31.17) | 1739 (48.64) | <0.001 |
|  | Low Risk | 1555 (84.69) | 281 (15.31) | 1836 (51.36) |  |

Obese males were nearly four times (AOR = 3.90; 95% CI: 2.12, 7.19) more likely to have hypertension compared to their normal counterparts while to obese females who were two-times (AOR =2.12; 95% CI: 1.55, 2.90) more likely to have hypertension compared to females with a normal BMI. Similarly, males with an abnormal waist circumference were nearly four times (AOR=3.78; 95% CI: 2.40, 6.03) likely to have hypertension while female participants were twice as likely. Both males (AOR=2.18; 95% CI: 1.70 2.80) and females (AOR =2.20; 95% CI: 1.64, 2.98) who had high risk WHtR had similar odds of having hypertension (Table 3).

**Table 3:**
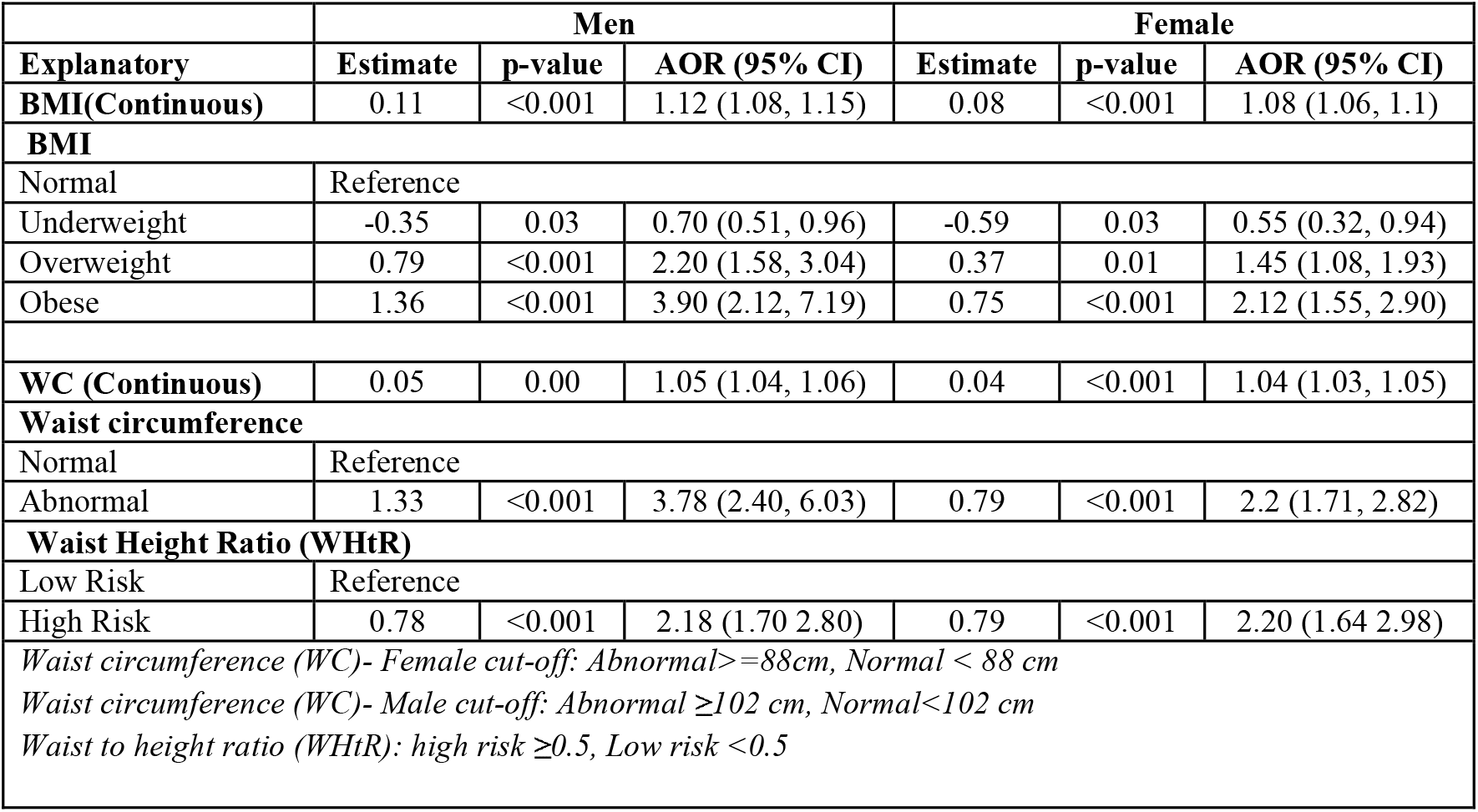
**Univariate Logistic regression between anthropometric measurements and its association hypertension**

Optimal waist circumference cut-off was 82.5cm for men and 87cm for women, with a sensitivity of 0.60 and 0.71 respectively. The WC cut-off for men was much lower than that recommended by the World Health Organisation (WHO) however, this study’s WC cut-off for women was similar to WHO recommendations. The optimal WHtR cut-off 0.47 and 0.55 for men and women respectively with both giving a similar sensitivity of 0.65. On the other hand, optimal BMI cut-off was 23.7 and 22.6 kg/m^2^ for male and females, respectively. The optimal BMI cut-off for men had the lowest sensitivity (0.39) compared to the other anthropometric measures for men; however, it still had a better sensitivity compared to the sensitivity of 0.06 obtained when WHO recommended cut-off for BMI among male individuals was used. The optimal BMI cut-off for females in this study gave the highest sensitivity of 0.78 compared to all other anthropometric measurements (Table 4).

**Table 4:** **Assessing optimal cut-off points for anthropometric predictors of hypertension**.

|  |  | Youden Method |  |  | WHO Recommended Cut-off s |  |  |  |
| --- | --- | --- | --- | --- | --- | --- | --- | --- |
|  | Optimal Cut-off | Sensitivity | Specificity | AUC | WHO Cut-off | Sensitivity | Specificity | AUC |
| **Waist circumference** |  |  |  |  |  |  |  |  |
| Men | 82.5 cm | 0.60(0.55,0.65) | 0.64(0.61,0.67) | 0.62 (0.59,0.65) | 102 cm | 0.13 (0.10-0.17) | 0.97(0.96-0.98) | 0.55(0.53-0.57) |
| Female | 87 cm | 0.71(0.66,0.75) | 0.55 (0.53, 0.58) | 0.63 (0.61,0.66) | 88 cm | 0.67 (0.62-0.71) | 0.59 (0.56-0.61) | 0.62(0.60 -0.65) |
| **Waist Height Ratio** |  |  |  |  |  |  |  |  |
| Men | 0.47 | 0.65(0.61, 0.70) | 0.60(0.57,0.62) | 0.62(0.59,0.65) | 0.50 | 0.49(0.44,0.54) | 0.75(0.73,0.78) | 0.62(0.59, 0.64) |
| Female | 0.55 | 0.65(0.60, 0.69) | 0.64(0.61,0.66) | 0.63(0.61, 0.67) | 0.50 | 0.83(0.79,0.86) | 0.39(0.36,0.41) | 0.61(0.58,0.63) |
| **BMI** |  |  |  |  |  |  |  |  |
| Men | 23.7 | 0.39(0.35-0.43) | 0.80(0.77-0.82) | 0.59(0.57-0.62) | 30 | 0.06(0.04-0.09) | 0.98(0.97-0.99) | 0.52(0.51-0.53) |
| Female | 22.6 | 0.78(0.74-0.812) | 0.39 (0.36-0.42) | 0.58(0.56-0.61) | 30 | 0.26(0.22-0.31) | 0.86(0.85-0.88) | 0.56(0.54-0.59) |

## Discussion

This study enrolled 3575 community members from four counties in Western Kenya with a mean age of 43.9 (±15.9) years of whom 1856 (51.6%) were female. This finding is slightly different from a community study conducted in Ethiopia [13] where the mean age of the study participants was 38.3 (±10.9) years, majority (54.2%) of whom were male. Although the difference in gender proportions in both studies was not statistically significant, the study conducted in Ethiopia [13] enrolled fewer (N=312) participants in a single (Kombolcha) town within Northeast Ethiopia compared to this larger community study that enrolled participants from a wider geographic region of Western Kenya. Although both the current study and that from Northeast Ethiopia [13] adopted the World Health Organisation (WHO) recommended STEPWISE tool to capture participants sociodemographic characteristics and nutritional status information; they still had differing community hypertension prevalence of 23.2% and 30.8% [13] respectively. Disease prevalence rates differ spatially and this could explain a higher community prevalence of hypertension in Ethiopia’s capital city (Addis Ababa) at 30.2% [12] and in Northwest Ethiopia at 28.3% [15]. In Nigeria’s Abia State, the prevalence of hypertension was found to be 31% [22] in a large survey (N=2928) that went house to house to determine the community prevalence of hypertension and its determinants. Similarly in Ghana’s community-based improvement program for hypertension [16], the prevalence of the disease was found to be 32.4% among 2337 individuals against a national average of between 20-32% [23].

This study reports a mean body mass index of 23.8 (±9.6) kg/m^2^ of which more than half (55%) of the community members enrolled had a normal BMI, 19.7% were overweight and 10.4% obese. From the findings of two studies conducted in Ethiopia [13,15], slightly higher mean BMI was reported at 25.7 (± 2.4) kg/m^2^ [13] and 24.7 (±4.6) kg/m^2^ [15]; a difference that could be attributed to a higher proportion (14.9%) of underweight participants enrolled in this study compared to 2.9% in Ethiopia [15]. In a study conducted in Ghana [16], the mean BMI index was 25.5 kg/m^2^ which is higher than the current study, a finding that could be attributed to lower proportion of underweight (7.3%) and normal (45.7%) participants. The likelihood of hypertension was higher among obese (AOR=3.9; 95% CI: 2.24, 6.81) male individuals compared to overweight (AOR=2.64.; 95% CI: 1.93, 3.58) males. The likelihood of obese females (AOR= 2.61; 95% CI: 1.96, 3.45) to be hypertensive was lower compared to their male counterparts. Although a study conducted in Zanzibar [24] did not stratify hypertension risk by gender rather than age categories, obese individuals older than forty years (AOR= 3.7; 95% CI: 1.3, 10.5) were more likely to be hypertensive compared to overweight individuals (AOR = 1.6; 95% CI: 0.7, 4.0) of the same age. In Saudi Arabia [25], obese individuals had a two-fold (OR= 2.22; 95% CI: 1.94, 2.54) increased likelihood of hypertension compared to those with a normal BMI. These two studies [24,25] confirm that a rise in body mass index is significantly associated with an increased risk of hypertension among the affected individuals.

This study further used the Younden method to identify optimal BMI cut-off for detecting hypertension. Among men, the optimal cut-off was 23.7 kg/m^2^ which was compared to the recommended WHO cut-off of 30 kg/m^2^ resulting in sensitivities of 0.39 (0.35-0.43) and 0.07(0.04-0.10) respectively. This signifies that with a lower BMI cut-off in rural Western Kenya, there is a higher the probability of detecting those with hypertension. However, a higher specificity of 0.98(0.97-0.99) was obtained using the WHO cut-off compared to 0.80 (0.77-0.82) that was obtained using the Younden cut-off for men. This study reports that to rule-out the presence of hypertension, it is appropriate to use the WHO recommended cut-offs. Overally, a BMI cut-off of 23.7 **k**g/m^2^ had a higher area under the curve (AUC) of 0.59(0.57-0.62) compared to 0.52 (0.51-0.53) when a cut-off of 30 kg/m^2^ was used. The AUC findings using a BMI cut-off of 23.7 kg/m^2^ compares to that in Korea [7] at 0.601. Similarly, in a second study conducted in Korea [26] that used nearly similar BMI cut-offs (23.59 kg/m^2^), the AUC was 0.58 (0.56, 0.60). However, in a study conducted in Nigeria [5], the AUC obtained using a Younden cut-off of 24.49 kg/m^2^ was 0.698 which is higher than the current study, with equally higher sensitivity of 0.73. However, these results from Nigeria reported lower specificity rate (0.60) than the current study, which could be attributed to the use of different BMI cut-offs. Overally, BMI is a poor predictor of hypertension among men living in rural western Kenya communities.

Among females the optimal BMI cut-off obtained was 27.1 kg/m^2^ which gave a sensitivity, specificity and AUC of 0.45(0.41-0.51), 0.72(0.70-0.74) and 0.58(056-0.61) respectively compared to the WHO recommended cut-off of 30 kg/m^2^ which had lower sensitivity (0.28) and AUC (0.56) but higher specificity (0.86). The use of BMI as a predictor of hypertension among women living in peri-urban and rural communities within Western Kenya is similar to studies conducted in Japan [8] and Cuba [18] which reported BMI as a good hypertension predictor for women. In a study targeting the Filipino-American women population living in four cities within the United States of America [27], a BMI cut-off of 25.5 **k**g/m^2^ was used giving higher sensitivity (0.65) and AUC (0.66) for detecting hypertension compared to the current study. A difference that could be attributed to average morphometric differences across race and geographic regions. Similarly among Korean women [26], when a BMI cut-off of 25.63 **k**g/m^2^ was used to predict hypertension, nearly similar sensitivity (0.43), specificity (0.71) and AUC (0.57) to the current study were obtained. These findings confirm the recommendations of previous studies [28,29] on the need for using region-specific BMI cut-offs to predict hypertension.

The mean waist circumference of the study participants was 85.37 (±12.26) cm which was comparable to the finding in Nigeria at 83.7 (±11.4) cm [22]. This study used similar waist circumference cut-off as those used in studies conducted in Ethiopia [13] and Nigeria [22] at 88cm for women and 102 cm for men. Nearly equal proportions of abnormal waist circumference were reported in this study (26.8%) and in Ethiopia (29.5%) [13] while lower proportions (20.7%) were noted in Nigeria [22]. We report that waist circumference is more sensitive for women (0.71) compared to men (0.60). This is similar to findings reported in Taiwan [7] where waist circumference alone was a good predictor of hypertension.

Lastly, the sensitivity and area under the ROC curve for waist height ratio was similar for both the male and female participants. This signifies that waist to height ratio alone can be used as a predictor of hypertension in western Kenya. Previous community screening programmes have over-relied on the use of BMI, however this study makes a case for the use of waist to height ratio which is both cheaper and less cumbersome. Furthermore, the sensitivity of waist to height ratio was highest for men in comparison to other anthropometric indicators. This is because waist to height ratio takes into account the individual’s body fat as well as their abdominal and central obesity which could be missed out when using BMI alone [5]. This study was conducted in peri-urban and rural western Kenya populations which are leaner compared to urban dwelling individuals in both Western Kenya and the rest of the country. This therefore implies that the findings of the study are not representative of what has been previously reported in more urbanized settlements within Kenya.

## Conclusions and Recommendations

Waist circumference and waist to height ratio are good predictors of hypertension among adult individuals irrespective of their gender. Body mass index has the highest sensitivity for detecting hypertension among women but the least sensitivity for men. We recommend the combined use of all the three anthropometric measurements as predictors of hypertension in local community screening programmes for the disease. There is need to review the use of BMI alone as a hypertension predictor among men in Western Kenya. Community screening programmes for hypertension should also consider using other hypertension markers such as biochemical tests and nutritional status alongside anthropometric measurements. Future studies using similar anthropometric measurements should be conducted in more urban settlements of Kenya to demonstrate socioeconomic differences in anthropometric cut-offs for hypertension.

## Data Availability

Data used in this study will be made publicly available on demand.

## Acknowledgements

This study received financial support from Access Accelerated through a cooperative agreement between World Bank and the division of non-communicable diseases (NCDs) in the Ministry of Health-Kenya. We would like to thank the management of Moi Teaching and Referral Hospital (MTRH), Moi University, Busia, Siaya, Vihiga and Trans Nzoia counties for offering an implementation agreement that enabled data collection in the respective counties. We are also grateful to the Academic Model Providing Access to Healthcare (AMPATH) for providing technical support for this study as well as the study participants for being part of this.

